# Patient Characteristics in Cases of Reinfection or Prolonged viral shedding in SARS-CoV-2

**DOI:** 10.1101/2021.05.14.21257231

**Authors:** Richard M. Yoo, Roland A. Romero, Joseph Mabajen, Suchit Mehrotra, Isaac S. Kohane, Natalie E Sheils

## Abstract

**Importance:** As testing options increase for COVID-19, their interpretability is challenged by the increasing variety of clinical contexts in which results are obtained. In particular, positive COVID-19 diagnostic (RT-PCR) tests that occur after a patient has seroconverted may be indicative of reinfection. However, in the absence of SARS-CoV-2 sequence data, the possibility of prolonged viral shedding may not be excluded. We highlight a testing pattern that identifies such cases and study its statistical power in identifying potential reinfection. We also study the medical records of patients that matched the pattern.

**Objective:** To describe the frequency and demographic information of people with a testing pattern indicative of SARS-CoV-2 reinfection.

**Design:** We examined 4.2 million test results from a large national health insurer in the United States. Specifically, we identified the pattern of a positive RT-PCR test followed by a positive IgG test, again followed by a positive RT-PCR.

**Setting:** Data from outpatient laboratories across the United States was joined with claims data from a single large commercial insurer’s administrative claims database.

**Participants:** Study participants are those whose insurance, either commercial or Medicare, is provided by a single US based insurer.

**Exposures:** People who received at least two positive diagnostic tests via RT-PCR for SARS-Cov-2 separated by 42 or more days with at least one serological test (IgG) indicating the presence of antibodies between diagnostic tests.

**Main Outcomes and Measures:** Count and characteristics of people with the timeline of three tests as described in Exposures.

**Results:** We identified 79 patients who had two positive RT-PCR tests separated by more than six weeks, with a positive IgG test in between. These patients tended to be older than those COVID-19 patients without this pattern (median age 56 vs. 42), and they exhibited comorbidities typically attributed to a compromised immune system and heart disease.

**Conclusions and Relevance:** While the testing pattern alone was not sufficient to distinguish potential reinfection from prolonged viral shedding, we were able to identify common traits of the patients identified through the pattern.

## Introduction

As testing options increase for COVID-19, their interpretability is challenged by the increasing variety of clinical contexts in which results are obtained. Most challenging are positive COVID-19 viral tests that occur after seroconversion has been confirmed by antibody testing. Do these suggest reinfection, variations in clearance of viral nucleic acids, or deficiencies test performance? Despite growing evidence of immunity from reinfection^1^ quantifying exceptions to this immunity remains elusive. We focus here on sequences of test results that might reflect reinfection or prolonged viral shedding (RPVS).

## Methods

We used de-identified administrative claims for individuals enrolled in a commercial, Medicare Advantage, or Medicaid plan from a single large health insurance provider in the United States. We joined this claims database with an outpatient dataset of individuals tested for SARS-CoV-2. These were linked with a daily record of patients admitted to a hospital with a primary or secondary diagnosis of COVID-19.

Our study population included all individuals with positive RT-PCR results. We excluded patients who had a negative RT-PCR test result reported the same day, had less than two positive RT-PCR tests separated by at least 42 days (to account for the typical length of viral shedding), and did not have a positive IgG test between two positive RT-PCR results.

With three consecutive positive tests in the specific pattern of: RT-PCR, IgG, RT-PCR there are multiple opportunities for false positives. To find the likelihood of a false positive across the RPVS subpopulation we compute the positive predictive value of a RT-PCR (sensitivity: 98%; specificity: 99.5%) and IgG test (sensitivity: 91%; specificity: 99.5%) using prevalence values computed from current New York Times reports^2^ and a likely ratio of estimated infections to reported case counts of 11 as suggested by the CDC.^3^

## Results

Of 234,866 patients that had positive RT-PCR results, 79 patients (0.034%) had two positive RT-PCR tests separated by more than six weeks, with a positive IgG test in between. Figure 1 shows the clinical timeline for those individuals.

**Figure 1:**
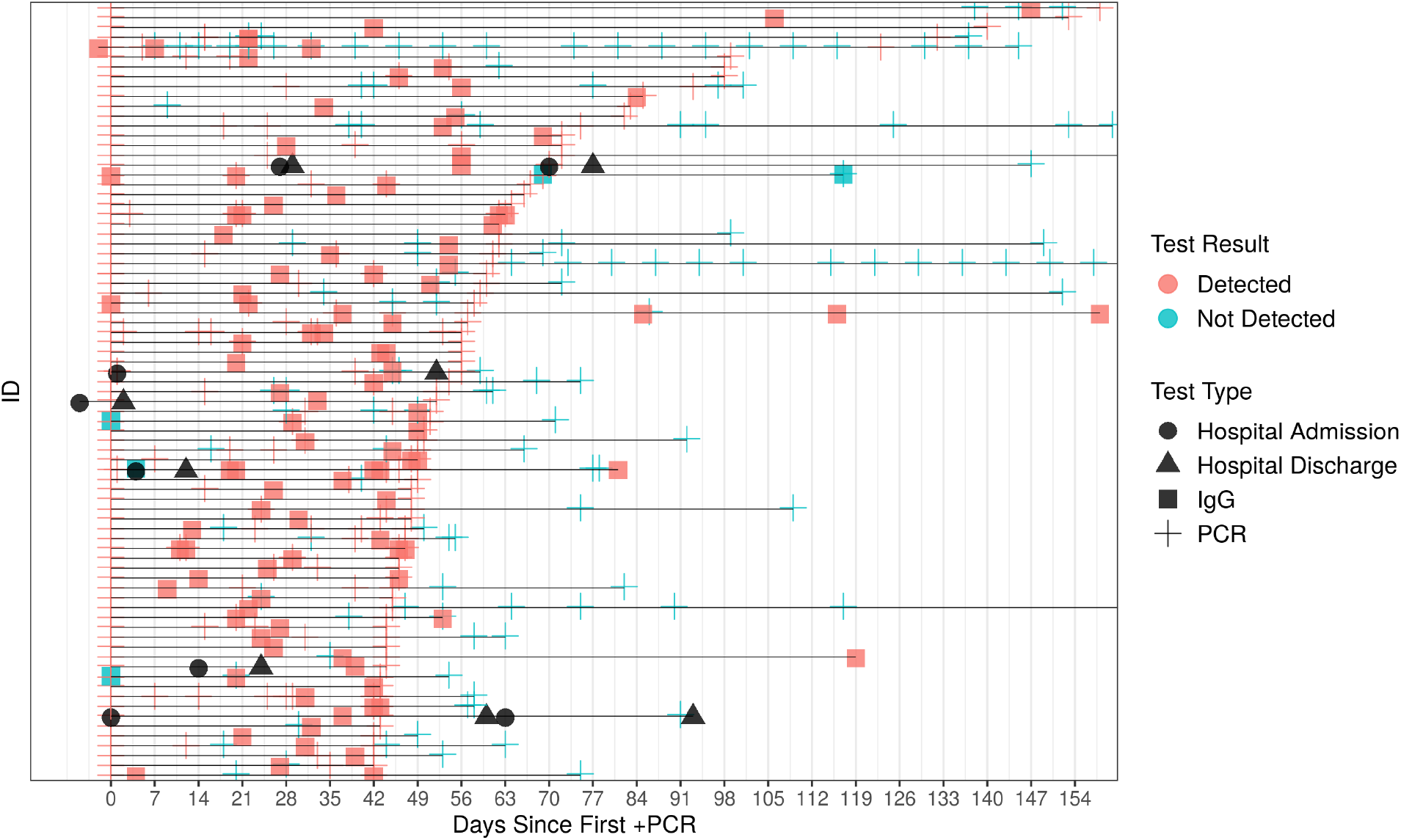
Timeline of tests for individuals with potential reinfection or prolonged viral shedding. That is, patients with the testing pattern, positive PCR, positive IgG, positive PCR with the two positive PCR tests separated by at least 42 days.

There are five possible combinations of false positives that would preclude RPVS in this population. The likelihood of one of these five occurring is 26.7%. Nonetheless, many of the individuals we consider have more than just three tests.

Because we could not recontact the individuals to resequence the virus, we cannot confirm reinfection. In the study population, the median number days between a positive IgG test and a subsequent positive RT-PCR test is 21 (IQR 24.5). Overall, a positive RT-PCR test could follow positive antibody test as quickly as one day later. Because of the ambiguity, we refer to these cases as putative or potential reinfection or prolonged viral shedding (RPVS) cases.

Figure 2 shows that comorbid conditions associated with a compromised immune system rank high on the list for patients with putative RPVS. The figure also shows the frequency of medical conditions for all the patients testing positive at least once for COVID-19. RPVS patients exhibit a higher frequency of comorbidities related to a compromised immune system than the general population who have a positive PCR test.

**Figure 2:**
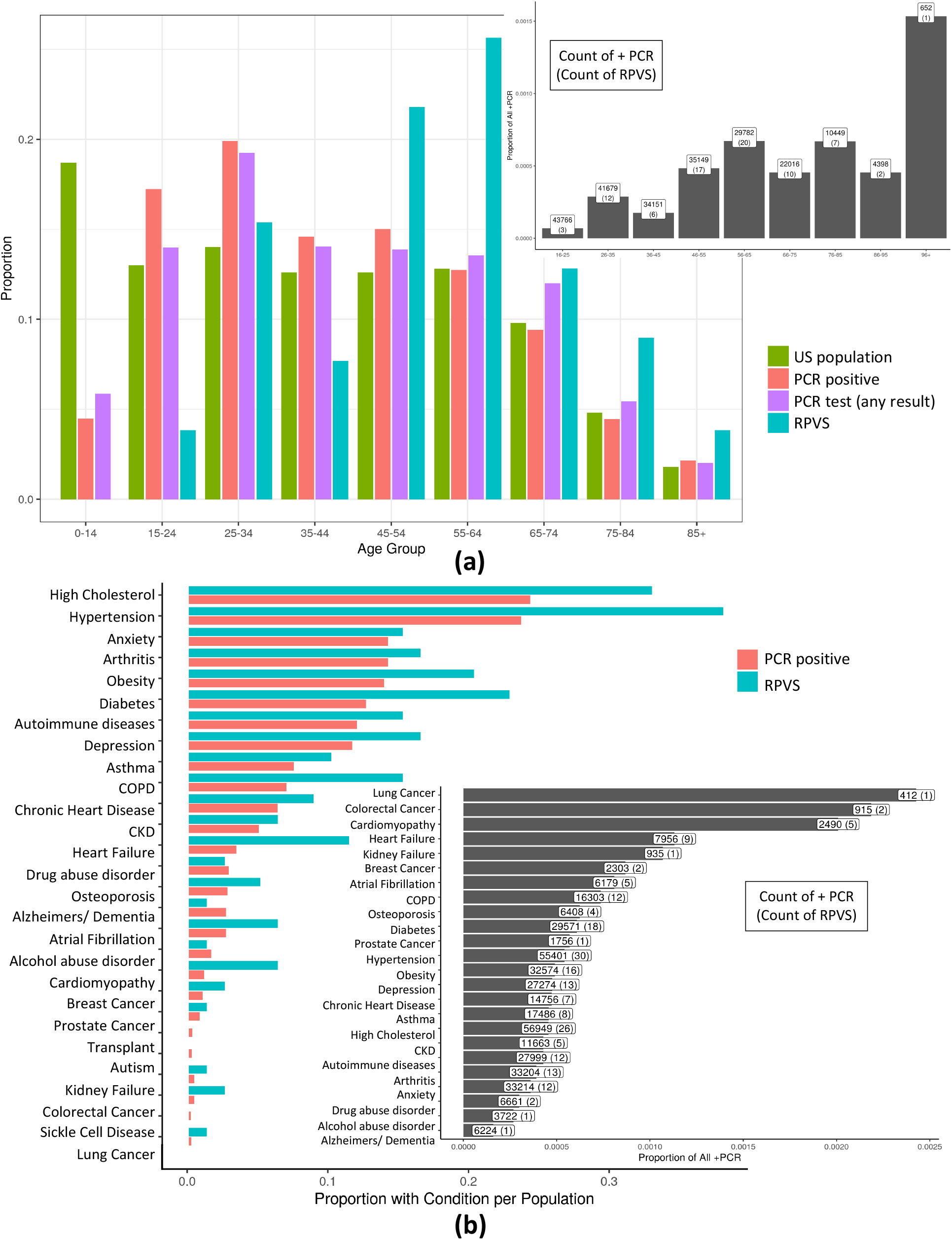
**a)** A normalized histogram of the US population, all those with a PCR test result in our database, those with a positive PCR test result, and suspected reinfection or prolonged viral shedding cases across patient age. For RPVS patients the median age is 56, and average is 54.65. In particular, 75% of cases are from age group above 44 suggesting a possible increased susceptibility to an RPVS state with age. Of note, this advanced age is not due to an ascertainment bias due to testing frequency with age since the highest group for per capita testing is aged 25-34. Further, RT-PCR positive cases (median = 42, average = 43.46) is shifted towards the younger ages relative to the putative RPVS cases. Further, the inset shows RPVS cases normalized by PCR positive cases in each age group. **b)** Histogram of comorbid conditions of those with COVID-19 (PCR positive) and RPVS patients. Again, the inset shows RPVS cases normalized by PCR positive cases for each comorbid condition

## Discussion

We present here cases of possible SARS-CoV-2 reinfection based on patterns of positive RT-PCR and positive antibody tests. We quantify the possibility of multiple false positive tests using testing accuracy reports and do not find these large enough to account for the findings. Also, reports have been made of prolonged viral clearance that could appear as reinfection in sick^4^ and “recovered” patients^5,6^ although the durations in those studies are considerably less than the cases we document here and we do not restrict to hospitalized patients.

## Data Availability

Data is not sharable due to patient privacy concerns.

